# Exploring the impact of parity and its interaction with history of preterm delivery on gestational duration

**DOI:** 10.1101/2023.02.16.23286023

**Authors:** Karin Ytterberg, Bo Jacobsson, Christopher Flatley, Julius Juodakis, Staffan Nilsson, Pol Sole-Navais

## Abstract

Delivering preterm is the leading cause of death in neonates and children under five years of age. Both genetics and environmental factors play a role in timing of delivery, and these influences can be unique to a single pregnancy or shared across pregnancies of the same mother. The aim of this study was to understand how gestational duration is affected by parity and how parity modifies the association between history of preterm delivery and gestational duration. To investigate this, we analysed 1 118 318 spontaneous deliveries (1990 - 2012) from the Swedish Medical Birth Register, with access to pedigrees, using linear regressions and linear mixed models. We found that parity has a modest effect on the mean and a large effect on the variance of gestational duration. Interactions with a woman’s clinical and family history of preterm delivery revealed both pregnancy-specific and shared factors. For instance, the effect of a previous preterm delivery on gestational duration is present across pregnancies, but the magnitude of its effect is pregnancy specific. The access to pedigrees made it possible to apply linear mixed models, thus including all woman’s pregnancies in the model and accounting for unobserved mother-specific covariates. The linear mixed models highlighted a group effect bias when using linear regression to estimate the association between parity and gestational duration, likely caused by socioeconomic factors. Our study shed light on how parity affects gestational duration and modifies the effect of well-known risk factors of preterm delivery.

## Introduction

Gestational duration, as well as preterm delivery (birth before 37 complete weeks of gestation), is a complex trait yet to be fully understood, making it difficult to prevent preterm birth. Preterm birth is the leading cause of death in neonates and children under five years of age [1]. The processes of the timing of delivery and preterm delivery are influenced by both genetic and environmental factors and can be either specific to a pregnancy or shared across pregnancies of the same mother. Specific factors are those acting on a specific pregnancy, while shared factors act with similar effects throughout more than one of a mothers’ pregnancies.

Parity, the number of times a woman has given birth to a child, is itself a well-known risk factor associated with preterm delivery. A woman carrying her first child has an increased risk of preterm delivery compared to women carrying their second child [2-5]. The risk of preterm delivery slightly increases again in the fifth pregnancy, but the risk is still lower than in the first pregnancy [3, 6]. However, the effect of parity on gestational duration, and whether it modifies the effect of known risk factors on gestational duration are not as well investigated.

A previous preterm delivery is one of the most relevant risk factors of preterm delivery [2, 4, 7], with the risk increasing for each additional previous preterm delivery [8-10]. The number of pregnancies between the preterm delivery and the current pregnancy also affects a woman’s risk of a preterm delivery [8-10]. Thus, a woman’s clinical history of preterm delivery could be an example of a specific factor affecting the risk of preterm delivery, and depends on when the preterm delivery occurred. Nevertheless, whether parity modifies the effect of a mother’s history of preterm births remains indeterminate.

Preterm delivery runs within families. Genome-wide-association studies explain a small fraction of the timing of delivery [12, 13], while epidemiological studies have shown that up to 30% of the variability in gestational duration and preterm delivery is due to genetics [11]. For instance, a mother herself being born preterm increases the risk of delivering preterm [14-16] and maternal female relatives giving birth preterm also confers higher risk of preterm delivery [15-16]. However, the duration of gestation of a father has a weaker impact on his child’s gestational duration [14]. Whether parity modifies the effect of family history of preterm delivery has been previously investigated, but with unconvincing results [16]. The effect of a maternal sibling delivering preterm appears to be larger in nulliparous women than in multiparous, but with overlapping confidence intervals. The use of gestational duration as a continuous outcome, in a much larger sample might clarify this potential effect modification, and that of other measures of family history.

Most previous studies assessing risk factors related to gestational duration and preterm delivery evaluated the risks between mothers. With pedigrees and mixed models, it is possible to account for unobserved mother-specific covariates and more accurately capture the true effect of well-known risk factors. To better understand how pregnancy specific and shared factors between a woman’s pregnancies act on gestational duration, we will, with our family pedigrees, investigate the effects of parity on gestational duration, and how it modifies the effects of major risk factors of preterm delivery with both linear and linear mixed models. We aim to answer 1) how parity affects the distribution of gestational duration, and 2) if parity modifies the effects of a woman’s clinical and family history of preterm delivery on gestational duration.

## Methods

### Sample selection

This study used data from three high-quality Swedish population-based registries: the Swedish Medical Birth Register, the Multi-Generation Register and the Education Register. The Swedish Medical Birth Register contains more than 96% of all births in Sweden with detailed information such as complications during pregnancy, neonatal period, delivery, reproductive history and demographic information [17-18]. Mother-child pairs in the Swedish Medical Birth Register covered the years between 1973 and 2012. Fathers for mother-child pairs were identified with the Multi-Generation Register [19]. Lastly, the highest level of completed education, between the years 1990-2012, was extracted from the Education Register [20] for mothers and fathers.

The pregnancies from the Swedish Medical Birth Register were stepwise filtered according to the flow chart in Supplementary Figure 1. Included pregnancies in the analysis were spontaneous singleton live births. Pregnancies with no maternal identification number, pregnancy duplications and pregnancies with no gestational duration were excluded. The data cleaning for outliers based on deviating child weight and gestational duration correlation was performed with a Generalised Additive Models for Location Scale and Shape (GAMLSS) [21] (Supplementary Methods 1). Filtering of assisted reproductive technology was performed on existing information in the Swedish Medical Birth Register and excluded pregnancies with action on infertility treatment, ovarian stimulation, surgery, intracytoplasmic sperm or other activity during infertility.

To be included as an index pregnancy the initiation of delivery had to be spontaneous. The spontaneous onset has only been recorded since 1990 in the Swedish Medical Birth Register, which dictated the exclusion of all pregnancies prior to 1990 and iatrogenic deliveries thereafter. Pregnancies with missing information on maternal age, maternal or paternal highest educational level, sex of the child, year of birth or mother’s birth country were also removed. To be included in any of the analyses the mother needed to have at least two spontaneous live births in the Swedish Medical Birth Register - note that this does not explicitly need to be the first and second pregnancies the mother delivered, it can be any two pregnancies existing in the available data.

### Variable definitions

In this study the “best estimate” of gestational duration according to the Swedish Medical Birth Register was used [18], which is based on either ultrasound or the latest menstrual period. Pregnancies ending before 259 days (37 weeks) were considered preterm deliveries.

Parity is the number of times a woman has given birth to a child, live or stillborn with a gestational duration ≥ 154 days (22 weeks). From 2008 and onwards, stillbirths were included in the Swedish Medical Birth Register if the duration of gestation was ≥ 154 days (the same as livebirths). Before 2008, stillbirths were included in the Medical Birth Register for gestations lasting ≥ 196 days (28 weeks). The counting of parity is zero-based, meaning that the first pregnancy is parity zero (nulliparous). Following this definition, the second pregnancy is counted as parity one and so on.

Preterm deliveries defining a woman’s clinical or family history of preterm delivery were not required to be spontaneous (Figure 1d), due to the lack of such data before 1990. The maternal clinical history of preterm delivery indicator variables were defined as follows:

**Fig. 1.**
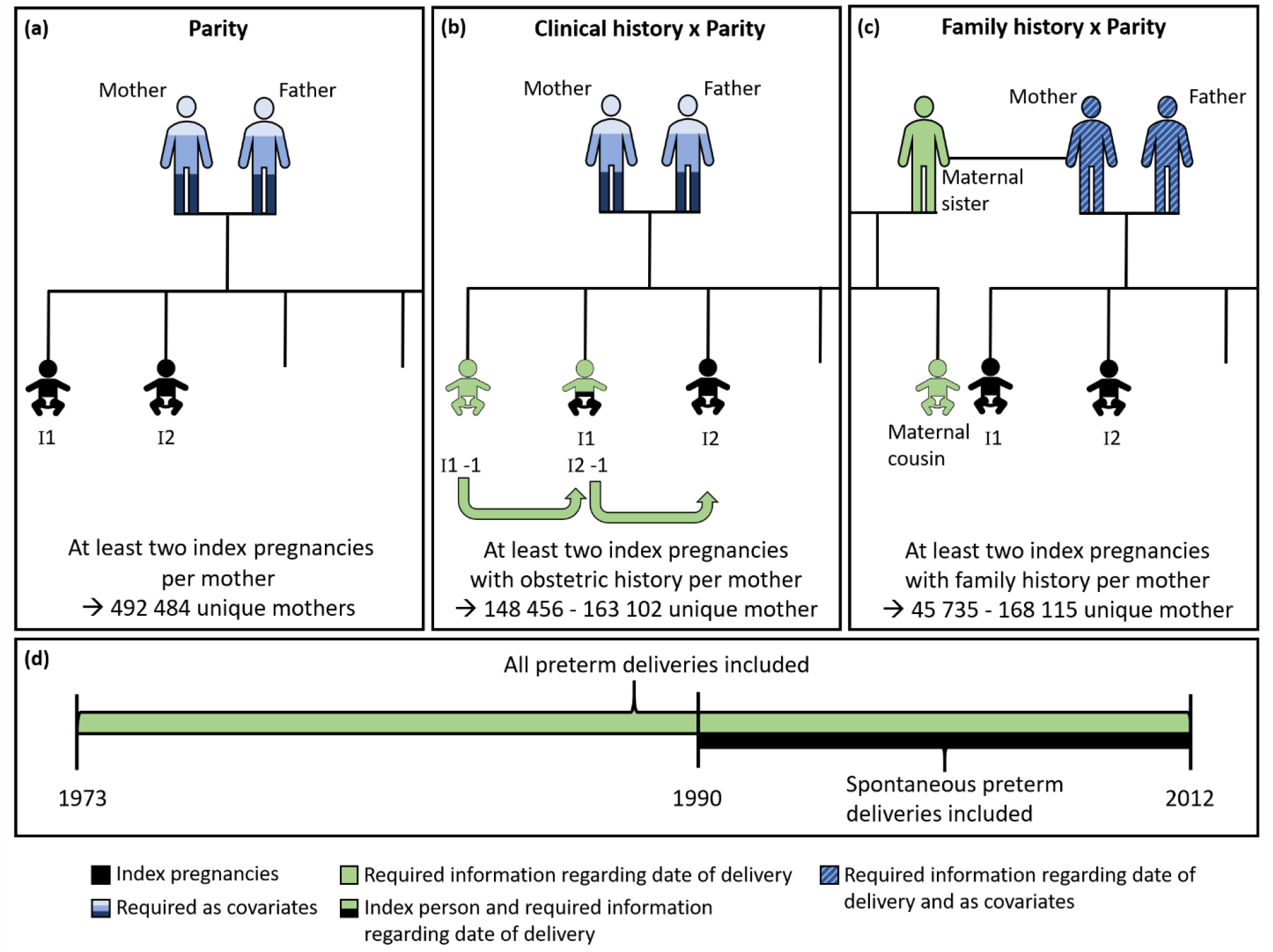
A diagram describing the family information required to perform the statistical analyses. This study evaluates (a) the association between parity and gestational duration, (b) the interaction between parity and clinical history of preterm delivery on gestational duration, and (c) the interaction between parity and family history of preterm delivery on gestational duration. Index pregnancies are indicated with an “I”, and to be included in any analysis at least two pregnancies from each mother were required. Information from mothers and fathers was required in all the analyses in (a), (b) and (c). When investigating the interaction between parity and clinical history of preterm delivery in (b), information regarding the gestational duration of the immediate previous pregnancy from the mother was needed. In (c), information regarding the mother’s, father’s, or maternal sister’s gestation duration, or the gestational duration of maternal sisters’ pregnancies, was required to perform any of the corresponding family history analyses. When information of gestational duration was required from any relative to the index pregnancy in (b) or (c), the delivery was not required to be spontaneous, as was for the index pregnancies (d).

- Previous preterm delivery: the nearest preceding pregnancy was preterm.
- Firstborn preterm delivery: the first pregnancy (parity zero) was preterm.

The family history indicator exposures were defined as:

- Mother born preterm: the mother of the index pregnancy was born preterm.
- Maternal sister born preterm: a maternal sister was born preterm.
- Father born preterm: the father of the index pregnancy was born preterm.
- Maternal sister given birth preterm: the maternal sister has herself delivered preterm (maternal cousin born preterm).

All information regarding family history was limited to the mothers/fathers/maternal sisters who are registered as a child in the Swedish Medical Birth Register.

### Statistical analyses

Linear regressions and linear mixed models were applied in this study to examine how gestational duration is affected by parity, and if parity modifies the effect of clinical or family history of preterm delivery on gestational duration. Mothers with only one pregnancy were not included in the study due to convergence issues in the linear mixed models. However, the results obtained when including all mothers were similar to those presented here.

To evaluate the association between parity and gestational duration, linear and linear mixed models were applied with three different sets of covariates. Set 1 of covariates adjusted for parity, sex and year of birth (linear and squared). Additional covariates were included in Set 2: Set 1 covariates, maternal birth country (Nordic vs. other), maternal age (in years, linear and squared), highest maternal education and highest paternal education (categorized in 7 ordered categories, SCB SUN 2000 categories). A third set of covariates, Set 3, included Set 2 covariates, unwilling subfertility, BMI (linear and squared), smoking, diabetes and preeclampsia. Note that the variables added in Set 3 were not available in as many pregnancies as those in Set 1 or Set 2, resulting in smaller sample size. The information required for each index pregnancy in this analysis is summarized in Figure 1a. For the linear models one random pregnancy per mother was selected and for the linear mixed models all pregnancies of a mother were included. Mixed models included a random effect for each mother, accounting for differences in mean gestational duration between mothers. Accounting for unobserved mother-specific covariates, allowed us to include multiple pregnancies per mother.

The variance and distribution of gestational duration in each parity was investigated by visual inspection of the distribution of gestational duration, estimation of the variance and assessment of significant differences in variance and distribution (Levene’s test and Kolmogorov-Smirnov test) by parity.

The effects of clinical and family history of preterm delivery, and their modification by parity, were investigated with similar models. First, the effect on gestational duration by each clinical and family history of preterm delivery exposure, described in “Variable definitions”, were assessed with linear mixed models. Then, the interaction between parity and each exposure variable was tested by adding an interaction term in separate models. Significant interactions (p<0.001, conservative significance level due to large sample size and the likelihood to detect smaller effect is higher) were further evaluated by analysing the effect of clinical or family history of preterm delivery variables in each parity separately using linear regression. In addition to the clinical or family history of preterm delivery exposures, the included covariate set in all steps were Set 2. The information required for each index pregnancy in this analysis is summarized in Figure 1b for the clinical history of preterm delivery and Figure 1c for the family history of preterm delivery. For a delivery to be included as a clinical or family history of preterm delivery it does not need to be a spontaneous delivery (Figure 1d).

### Data access

Due to sensitive information, access to raw data requires an application to the National Board of Health and Welfare and Statistics Sweden, conditioned on an approved ethical application. All analyses were executed with Ubuntu 20.04.3 LTS and in R 4.0. Moreover, analysis workflow was managed using Snakemake [22], in a defined Conda environment [23]. All code is available [here](https://github.com/PerinatalLab/Parity_gestational_duration_interaction).

## Results

### Descriptive statistics

Description of study participants, mothers with two or more spontaneous live births, are shown in Table 1. The median of gestational duration in all pregnancies was 281 days and the highest incidence of spontaneous preterm birth was observed in the mothers’ first pregnancy (parity 0) at 4.6%, while the lowest incidence was observed in the second pregnancy at 2.7%. Overall, the occurrence of spontaneous preterm delivery was 3.5% for all included pregnancies. Any clinical or family history of preterm delivery was approximately equally prevalent across parities and the proportion of mothers and fathers born preterm were approximately the same. Moreover, it was more common that mothers were born in a Nordic country and that parents had at least an upper secondary school degree or higher.

**Table 1.** Baseline characteristics of 1 118 318 singleton spontaneous live births by parity in Swedish population^a^

| Variable | Parity |  |  |  |  |
| --- | --- | --- | --- | --- | --- |
|  | 0 | 1 | 2 | ≥3 | All |
| **Number (%)** | 418 147 (37) | 462 021 (41) | 169 453 (15) | 68 697 (6.1) | 1 118 318 (100) |
| **Gestational duration**, median (interquartile range) | 281 (274–287) | 281 (275–286) | 281 (275–286) | 281 (274–286) | 281 (275–286) |
| **Preterm delivery (%)** | 19 386 (4.6) | 12 498 (2.7) | 4 551 (2.7) | 2686 (3.9) | 39 121 (3.5) |
| **Sex Female (%)** | 204 422 (49) | 225 514 (49) | 83 396 (49) | 33 869 (49) | 547 201 (49) |
| **Year of birth**, median (interquartile range) | 1999 (1994–2005) | 2002 (1996–2007) | 2002 (1997–2007) | 2002 (1996–2007) | 2001 (1995–2006) |
| **Maternal age**, median (interquartile range) | 26 (23–29) | 29 (26–32) | 32 (28–35) | 34 (30–37) | 29 (25–32) |
| **Highest maternal education (%)** |  |  |  |  |  |
| < 9 years of schooling | 5 121 (1.2) | 6 764 (1.5) | 4 885 (2.9) | 5 768 (8.4) | 22 538 (2.0) |
| 9 years of schooling | 23 699 (5.7) | 27 722 (6.0) | 14 497 (8.6) | 10 814 (16) | 76 732 (6.9) |
| upper secondary ≤ 2 years | 72 611 (17) | 85 989 (19) | 39 694 (23) | 21 062 (31) | 219 356 (20) |
| upper secondary = 3 years | 108 409 (26) | 116 565 (25) | 38 032 (22) | 13 085 (19) | 276 091 (25) |
| Post-secondary < 3 years | 63 444 (15) | 69 537 (15) | 23 485 (14) | 7 126 (10) | 163 592 (15) |
| Post-secondary ≥ 3 years | 139 939 (33) | 150 155 (32) | 47 073 (28) | 10 488 (15) | 347 655 (31) |
| Postgraduate education | 4 924 (1.2) | 5 289 (1.1) | 1 787 (1.1) | 354 (0.51) | 12 354 (1.1) |
| **Highest paternal education (%)** |  |  |  |  |  |
| < 9 years of schooling | 5 563 (1.3) | 6 819 (1.5) | 4 213 (2.5) | 4 126 (6) | 20 721 (1.8) |
| 9 years of schooling | 39 996 (9.6) | 44 966 (9.7) | 19 802 (12) | 11 170 (16) | 115 934 (10) |
| upper secondary ≤ 2 years | 121 934 (29) | 137 372 (30) | 55 307 (33) | 25 719 (37) | 340 332 (30) |
| upper secondary = 3 years | 91 059 (22) | 98 540 (21) | 31 369 (19) | 10 953 (16) | 231 921 (21) |
| Post-secondary < 3 years | 63 206 (15) | 69 671 (15) | 23 631 (14) | 7 491 (11) | 163 999 (15) |
| Post-secondary ≥ 3 years | 88 519 (21) | 95 993 (21) | 31 838 (19) | 8 401 (12) | 224 751 (20) |
| Postgraduate education | 7 870 (1.9) | 8 660 (1.9) | 3 293 (1.9) | 837 (1.2) | 20 660 (1.8) |
| **Nordic country of maternal birth (%)** | 365 304 (87) | 400 809 (87) | 140 680 (83) | 50 146 (73) | 956 939 (86) |
| **Firstborn preterm (%)<sup>b,h</sup>** | 19 386 (4.6) | 21 696 (4.8) | 8 287 (5.4) | 3 359 (6.0) | 52 728 (4.9) |
| **Previous preterm (%)<sup>c,h</sup>** | - | 21 696 (4.8) | 5 205 (3.2) | 2 534 (3.8) | 29 435 (4.3) |
| **Mother preterm (%)<sup>d,h</sup>** | 6 169 (3.8) | 6 262 (3.8) | 1 698 (4.0) | 468 (4.9) | 14 597 (3.9) |
| **Maternal sister born preterm (%)<sup>e,h</sup>** | 1 612 (3.7) | 1643 (3.7) | 527 (4.1) | 176 (5.6) | 3 958 (3.8) |
| **Maternal sister given birth preterm (%)<sup>f,h</sup>** | 1 300 (3.0) | 2 017 (4.5) | 731 (5.7) | 271 (8.6) | 4 319 (4.1) |
| **Father preterm (%)<sup>g,h</sup>** | 5 006 (4.3) | 5 265 (4.4) | 1 406 (4.5) | 410 (5.4) | 12 087 (4.4) |

<sup>a</sup> The table captures the pregnancies' included in covariate Set 1 and Set 2. For description of the additional maternal covariates in Set 3 see Supplementary Table 1.
<sup>b</sup> Missing's: 41 389 pregnancies
<sup>c</sup> Missing's: 440 222 pregnancies
<sup>d</sup> Missing's: 741 217 pregnancies
<sup>e</sup> Missing's: 1 014 352 pregnancies
<sup>f</sup> Missing's: 844 406 pregnancies
<sup>g</sup> Missing's: 1 014 242 pregnancies
<sup>h</sup> History of preterm delivery includes both spontaneous and iatrogenic deliveries

### Parity affects the mean of gestational duration

The association between parity and gestational duration was evaluated with linear regression and linear mixed models (Figure 2). Results obtained with covariate Set 1 and Set 3 were similar to those from the models with Set 2, therefore only results from Set 2 will be presented further. Perhaps unsurprisingly, the mean gestational duration differed by parity. However, compared to the second pregnancy (the one with the lowest incidence of preterm delivery), the gestational duration in the first pregnancy was only 6-8 hours shorter on average (beta: -0.29 days; 95% CI: -0.33, -0.25 for the linear mixed regression Set 2). The gestation of the third pregnancy was longer compared to the second pregnancy, but again only a few hours on average (beta: 0.37 days; 95% CI: 0.31, 0.42 for linear mixed regression Set 2). For the fourth and subsequent pregnancies the gestational duration was shorter than the second pregnancy (beta: -0.13 days; 95% CI: -0.23, -0.04 for the linear mixed regression Set 2).

**Fig. 2.**
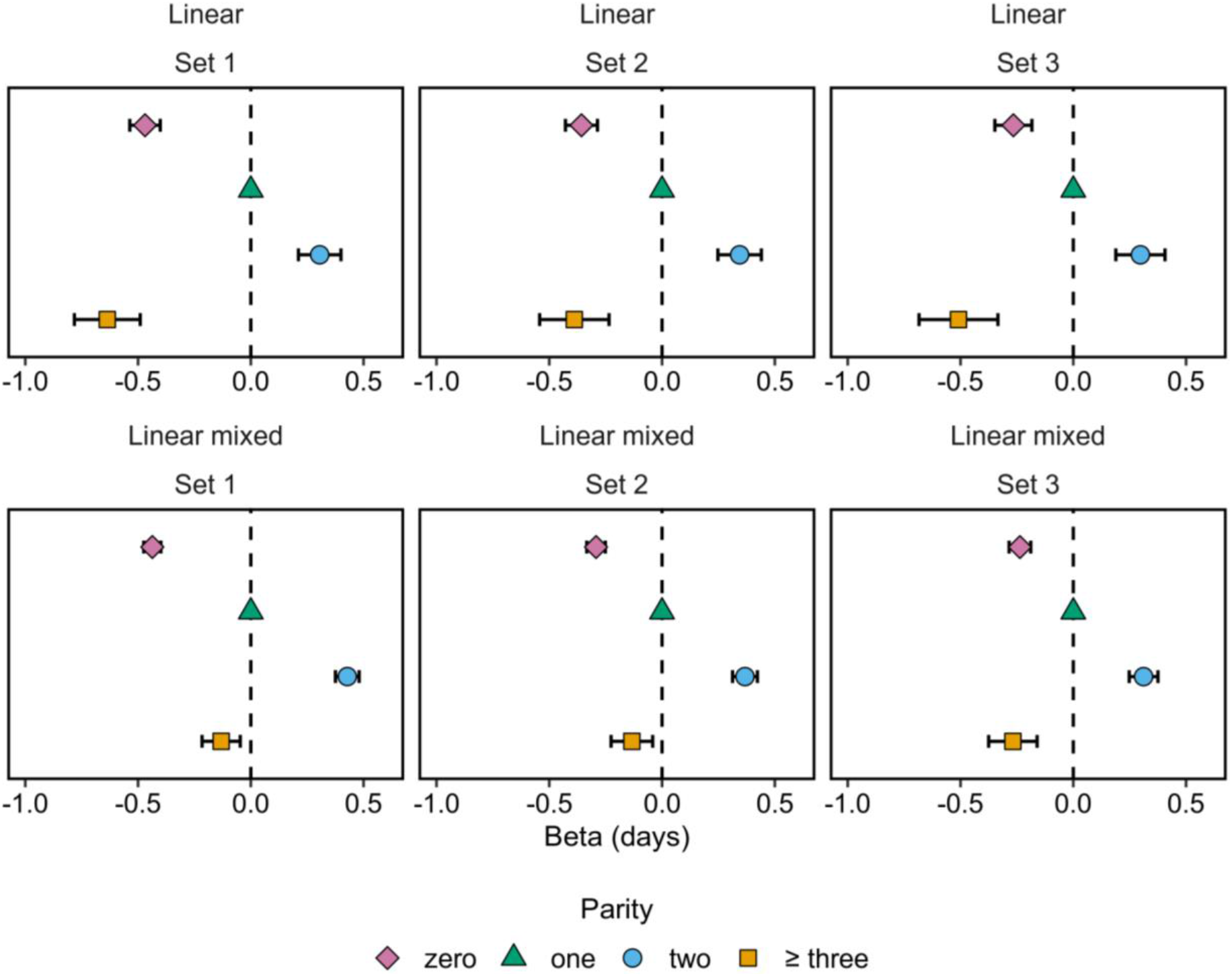
The association between gestational duration and parity in linear regression and linear mixed models. The reference group was the second pregnancy (triangle) and compared to the first (diamond), third pregnancy (circle) and fourth and subsequent pregnancies (square). In the linear regressions (top), one random pregnancy was selected for each mother with at least two pregnancies, resulting in a sample size of 492 484 pregnancies for Set 1 and Set 2, and a sample size of 360 343 for Set 3. While in the linear mixed models (bottom) all pregnancies of the mothers were included, with a random effect for the mother, resulting in a sample size of 1 118 318 pregnancies and 492 484 unique mothers for Set 1 and Set 2, and 801 843 pregnancies and 360 343 unique mothers for Set 3.

Notably, the magnitude of effect was larger for the fourth and subsequent pregnancies in the linear model compared to the linear mixed model (beta: -0.39 days; 95% CI: -0.54, -0.24 for linear regression Set 2). One hypothesis is that the total number of children a mother will have is associated with gestational duration, possibly due to maternal socio-economic factors. This would not affect the linear mixed models since they include a random effect to account for these, but will in the linear model cause bias due to sampling one random pregnancy per mother. This group effect bias was verified in a simulation (Supplementary Methods 2). The linear and linear mixed model only diverged when the simulated groups had both a different total number of pregnancies and a group effect (different average gestational duration) (Supplementary Table 2). When the groups had the same total number of children with the group effect, or if the groups had a different number of children but no group effect, the shift in estimate between the linear and mixed linear model did not appear. This is one explanation for the shift between the models in the estimate of parity effect observed in pregnancy four and later (Figure 2).

### Parity affects the variance of gestational duration

In the distribution of gestational duration by parity there were large differences in the variance across parity (Figure 3a, NB: unadjusted). The largest variance was observed in the first pregnancy, and the lowest in the second pregnancy (Levene’s test, p-value < 1 × 10^−15^). This difference in variance is caused by shifts in the tail of the distribution (Figure 3b). The quantile-quantile plot for gestational duration between the first and second pregnancy (Kolmogorov-Smirnov test, p-value < 1 × 10^−15^), indicated a larger variance in the first pregnancy due to more deliveries in the early preterm period - which likely reflects the higher PTD rates despite a very small difference in mean gestational duration (Figure 3b and Supplementary Table 3, NB: unadjusted).

**Fig. 3.**
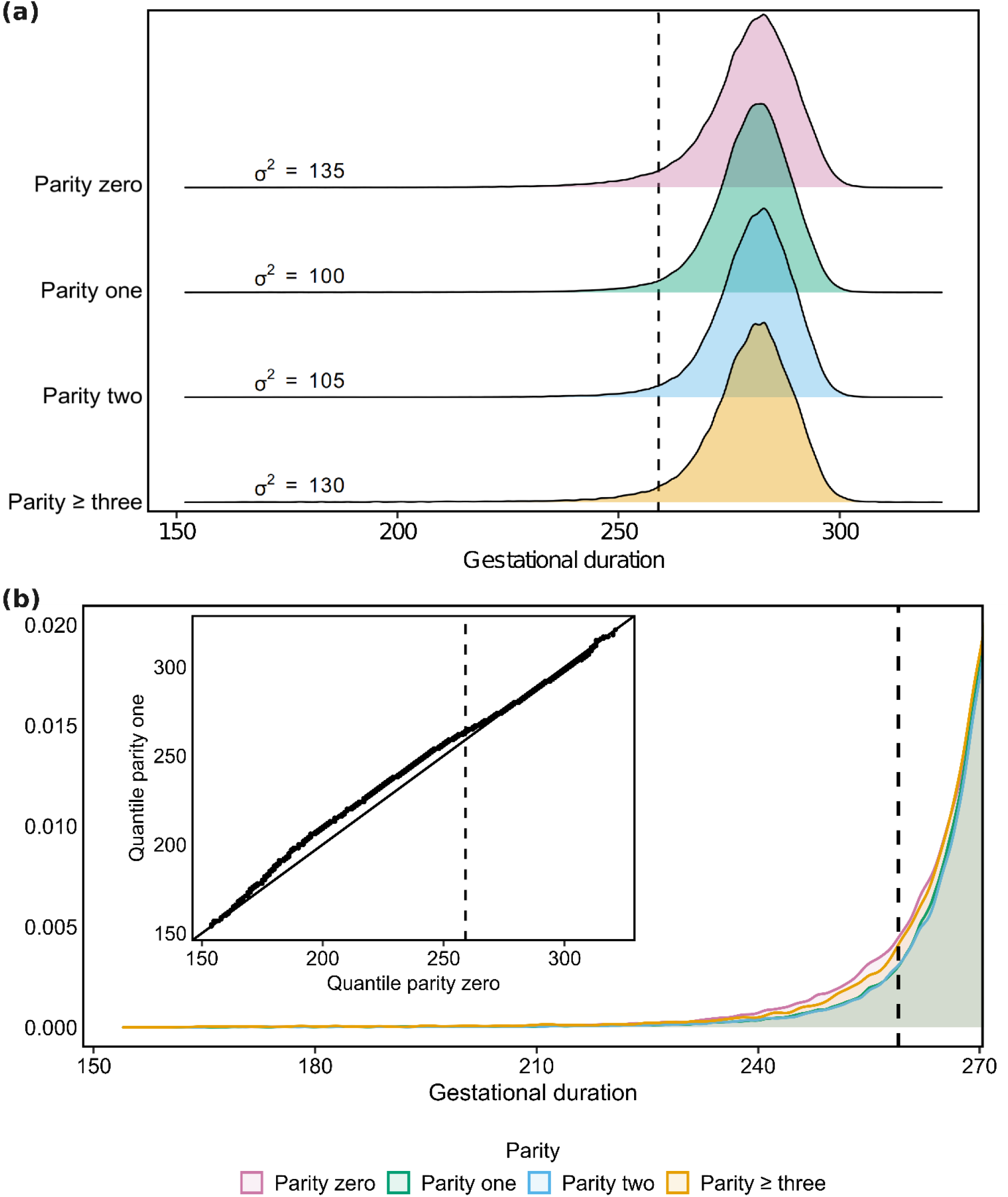
Distribution of gestational duration by parity. The dashed line represents the cut-off for a preterm pregnancy (259 days, 37 weeks). (a) Density plots of gestational duration by parity. All pregnancies were included in the analysis (parity zero 418 147 pregnancies; parity one 462 021 pregnancies; parity two 169 453 pregnancies; parity ≥ three 68 697 pregnancies). (b) Tail distribution of gestational duration by parity, truncated at 270 days. All pregnancies that were included in (a) were included in the tail distribution. The quantile-quantile plot of gestational duration in the first and second pregnancy (inset (b)) includes only mothers with a first and second-born. This resulted in 407 224 pregnancies in both parity zero and parity one, respectively.

### A clinical history of preterm delivery contributes to both pregnancy specific and shared factors

The modifying effect of a woman’s clinical history of preterm delivery on gestational duration by parity was then investigated. Both a preterm firstborn and previous (i.e. immediately preceding current) preterm delivery shorten the gestational duration of the index pregnancy (beta: -7.40 days, 95% CI: -7.58, -7.21; beta: -9.74 days, 95% CI: -9.91, -9.50 respectively, linear model with Set 2 and the specific clinical history of preterm delivery variable). Mixed models were not used here because the random effect is derived from the exposure, which would complicate the interpretation (since the exposure is the outcome of one of the mother’s pregnancies). When including an interaction term, the effect of both history variables was significantly modified by parity (p-values < 1× 10^−10^). The interaction was investigated by analysing the clinical history of preterm delivery measurements separately in each parity using linear regression (Figure 4). Note, that for the second pregnancy, the two history of preterm delivery measurements have an identical definition, and it is not possible to estimate these effects in a mother’s first pregnancy since such women do not have any previous deliveries. If the first pregnancy resulted in preterm delivery, the gestational duration was roughly eight days shorter (beta: -8.34 days; 95% CI: -8.58, -8.11) in the second pregnancy. In subsequent pregnancies, the effect of delivering a firstborn preterm was smaller, but still six and a half days shorter on average (beta: -6.68 days; 95% CI: -6.91, -6.46; beta: -6.67 days; 95% CI: -7.14, -6.19, in pregnancy three and four respectively). The patterns in pregnancy three and four could indicate that from pregnancy three, a firstborn preterm is a shared effect between the pregnancies shortening any future pregnancy with approximately 6.7 days. Moreover, a previous preterm birth in the third or fourth pregnancy was associated with a greater pregnancy length reduction compared to the second pregnancy (beta: -11.0 days; 95% CI: -11.3, -10.9; beta: -13.0; 95% CI: -13.6, -12.4, in parity two and three respectively). Note that the effect was larger in the fourth pregnancy than in the third, and so the history of a previous preterm birth acts as a pregnancy specific risk factor, with effect increasing in magnitude with each parity. A sensitivity analysis which only included clinical history of spontaneous preterm delivery showed similar results (Supplementary Figure 2). When the clinical history variables for all previous pregnancies were included in a single model, the pattern seen in Figure 4 still exists (Supplementary Tables 4 and 5), and the more recent the previous preterm delivery was, the larger the effect on gestational duration.

**Fig. 4.**
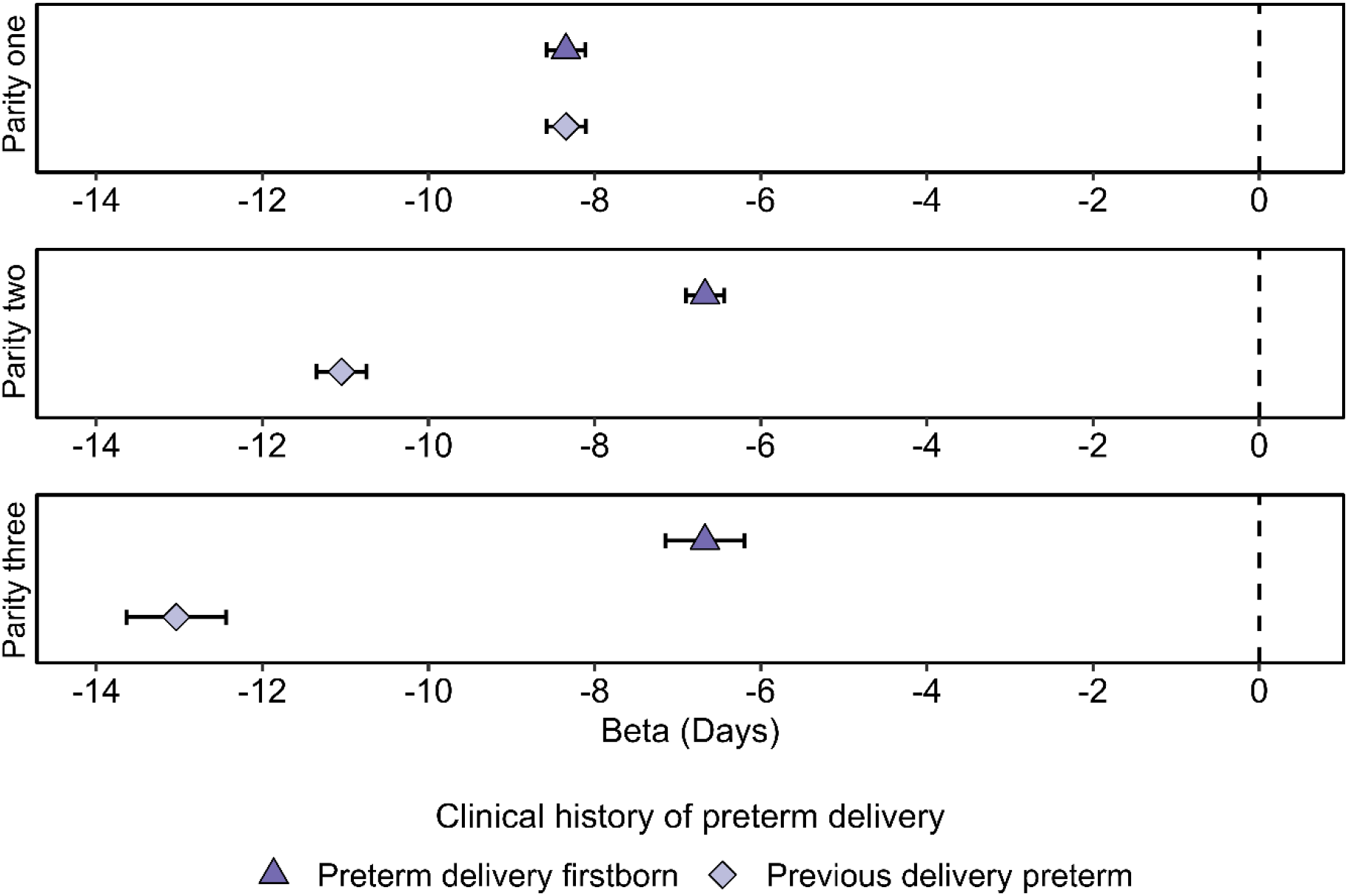
The modifying effect of a woman’s clinical history of preterm delivery on gestational duration by parity. The triangle represents the reduction of gestational duration in days if a firstborn was born preterm, while the diamond represents the reduction on gestational duration in days if the previous pregnancy was born preterm. For the second pregnancy, the two history of preterm delivery measurements have an identical definition. Linear models were run separately for each parity and the sample size for the different models were: Preterm delivery firstborn: parity one 135 406; parity two 144 092; parity three 35 985 and Previous delivery preterm: parity one 134 687; parity two 146 340; parity three 37 516. The linear models ran were adjusted for Set 2 and the specific clinical history of preterm delivery variable.

### Family history may have pregnant-specific effects in the first pregnancy

The effect of family history of preterm delivery on gestational duration (mother born preterm, maternal sister born preterm, father born preterm, and maternal sister given birth preterm) by parity was analysed with linear and linear mixed models. When not stratified by parity (Supplementary Figure 3), a mother herself born preterm, a maternal sister born preterm and maternal sister given birth preterm shortened gestational duration by approximately one and a half to two days (beta: -1.75 days; 95% CI: -1.96, -1.53; beta: -1.78 days; 95% CI: -2.20, -1.37; beta: -2.38 days; 95% CI: -2.70, -2.05 respectively in linear mixed models). When the father was born preterm the gestational duration decreased to half a day (beta: -0.63 days; 95%; CI: -0.86, -0.40, in a linear mixed model).

All four family history exposures were tested for interaction with parity on gestational duration. There was one significant interaction: a maternal sister delivering preterm (p-value = 4 × 10^−6^, all other tests p-value ≥ 0.01). Here the reduction in gestational duration was larger in the first pregnancy and reduced the gestational duration by approximately one more day compared to any other pregnancy (Figure 5). Furthermore, a three-way interaction between the mother’s own preterm birth status, parity and the parity of the mother when she was born, was investigated, with covariate Set 2. The interaction was tested in a linear mixed model and investigated parity-wise in linear regression. There was some evidence pointing towards that firstborn mothers, born preterm themselves, had a shorter gestational duration when carrying their first child, compared to if the mother was born preterm in any other pregnancy than the first (p-value = 0.009, Supplementary Figure 4).

**Fig. 5.**
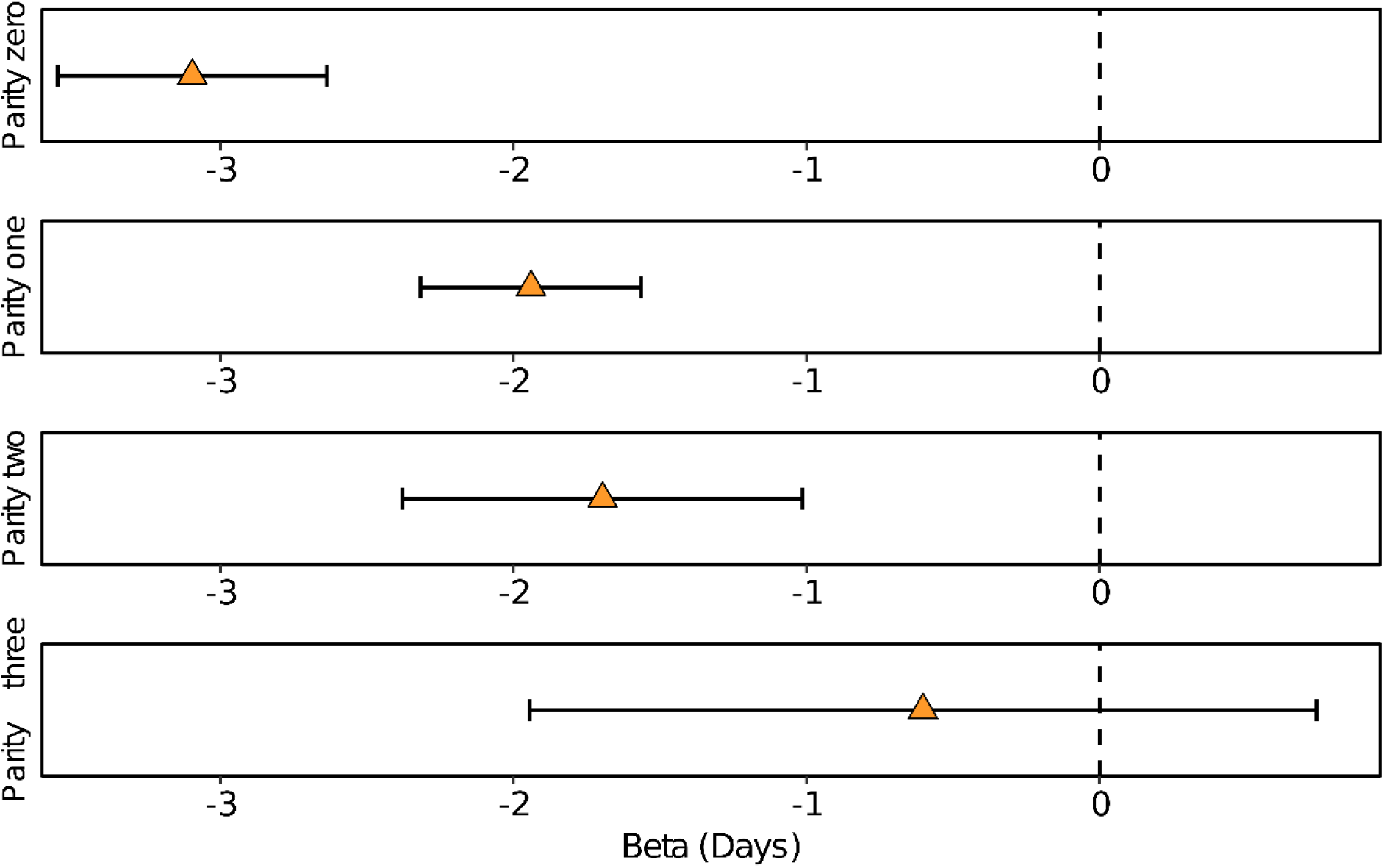
Parity modifies the effect on gestational duration when a maternal sister delivered at least one child preterm. The linear models were run separately for each parity with a sample size of parity zero: 51 295; parity one: 47 231; parity two: 13 499; parity three or later: 3 348. The models were adjusted for with Set 2 and the maternal sister given birth preterm exposure.

Overall, maternal sister delivering preterm is a pregnancy specific factor for the first pregnancy, and there is some evidence on additional pregnancy specific family history factors in the first pregnancy. In a mother’s later pregnancies family history variables act as shared factors.

### Parity has cohort effects on gestational duration

There was a significant interaction between parity and year of birth on gestational duration for the first and third pregnancy (p-value ≤ 1 × 10^−7^, with the second pregnancy as the reference). The interaction was not significant for the fourth and subsequent pregnancies (p-value ≥ 0.003). Overall, the mean gestational duration by parity varied approximately half a day over the years 1990 to 2012 (Supplementary Figure 5a). Year of birth modifies the effects of parity in both univariable and multivariable analyses (Supplementary Figure 5a and 5b), suggesting that these modifying effects may be caused by an external factor, for example changes in clinical procedures.

The effect of maternal age on gestational duration varies with parity in the first and third pregnancy (p-value ≤ 1 × 10^−5^, second pregnancy as reference) (Supplementary Figure 6). However, there is no variation in gestational duration for the fourth and subsequent pregnancies by maternal age (p-value > 0.7, second pregnancy as reference). For mothers younger than 20 years, there was no negative effect of parity zero: the mean gestational duration in first pregnancy was even longer than in the second (reference) pregnancy (beta = 0.75 days; 95% CI: 0.02, 1.49). In mothers above 40 years of age the mean gestational duration does not differ between parities. Mothers between the ages 20 and 39 had approximately the same pattern as in Figure 1, except that the pregnancy four and later was not significantly different from the second pregnancy.

## Discussion

In this paper, we investigated the effect of parity on gestational duration and assessed whether a woman’s clinical and family history of preterm delivery are pregnancy-specific or shared factors across pregnancies. We showed that parity has a modest effect on the mean, but a large effect on the variance of gestational duration, modifies the effect of clinical history of preterm delivery and the effect of a maternal sister delivering preterm.

Previous studies have shown that the first pregnancy is associated with an increased risk of preterm delivery compared to the second pregnancy [2-5]. Our results show the same pattern with a shorter mean gestation in the first pregnancy, but with a small effect (6-8 hours). Interestingly, we also observed that the variance of gestational duration is the largest in the first pregnancy due to more deliveries occurring in the preterm period. The effect of parity on average gestational duration is clinically negligible (a few hours), but the large difference in variance points towards a particular relevance for preterm deliveries, indicating time-varying effects acting in nulliparous women.

The gestational duration is shorter in mothers with a firstborn delivered preterm or a previous preterm delivery, which is consistent with previous research [2, 4, 7]. We also know from previous research that the closer the current pregnancy is to the preterm delivery, the larger the effect in determining gestational duration [8-10]. However, the reduction in gestational duration increased with increasing parity for a previous preterm delivery, which suggests the effect is shared across pregnancies, but the magnitude of effect is pregnancy specific. Also, a firstborn preterm seems to have the same effect on subsequent pregnancies, which can be seen as a shared effect between these pregnancies. Overall, we believe that the effect of clinical history of preterm delivery on gestational duration is not a direct causative effect, instead it captures maternal characteristics/effects that are shared across pregnancies.

The biological explanations for the effect on gestational duration by parity and clinical history of preterm delivery can only be speculated. For instance, immune response is associated with preterm delivery [24-25] and could be one mechanism causing distinct physiology of preterm delivery in different parities [26-28]. Furthermore, immunological memory could characterize a future pregnancy based on the outcome of previous pregnancies, which have been found to affect subsequent pregnancy for a woman with preeclampsia [29]. Parity needs to be further investigated to understand its biological mechanisms and effect.

If we assume that family history is a proxy for genetic factors which contribute equally in each parity (no interaction with mother herself born preterm), then the phenotypic variance explained by the environment will be larger in the first pregnancy, since the variance of gestational duration is larger in the first pregnancy compared to the second. An equal amount of genetics but a different contribution from environmental factors would result in different heritability estimates of gestational duration by parity (with the lowest in the first pregnancy). In contrast with this, we observed some evidence that firstborn mothers born preterm themselves had a lower gestational duration when carrying their first child compared to if the mother was born preterm in any other pregnancy but the first. A modifying effect with parity was seen for when a maternal sister had delivered at least one child preterm (confirm previous research [16]). This could be explained by some pregnancy-specific factor affecting a mothers first pregnancy and may have implications in the design of genetic studies attempting to discover genetic variants associated with gestational duration or preterm delivery. However, both the explanation regarding heritability and pregnancy-specific factors are currently speculations, and a formal test on genetic data should be employed to confirm.

We cannot rule out unmeasured confounders that are not shared between pregnancies of a mother, and these would not be captured by linear mixed models. Another factor of the Swedish population is that its prenatal care is advanced, comprehensive and available to all inhabitants. Similar analyses should therefore be performed in various populations to ensure robustness in the results presented here. A methodological aspect to highlight given the difference in variance of gestational duration by parity is the assumption of residuals having constant variance in linear regression. Since the variance of gestational duration is not constant over parity, the residual variance will not be constant. This will affect the size of the p-values, but not the estimates of the model. Choosing models which incorporate such variance differences when investigating the effects of gestational duration by parity could be interesting to explore. Another aspect to highlight is that maternal characteristics may be correlated to the total number of children the mother has carried. This was seen when comparing the high-parity effect estimates of the linear and linear mixed model where the magnitude of effect on gestational duration was lower in the latter. This is an important aspect in the choice of method (linear mixed models more unbiased than a linear model), but also highlights an important aspect of what parity captures.

This study hopes to bring a better understanding of how some of the most relevant known risk factors of preterm delivery act on gestational duration across a woman’s pregnancies. To conclude, we have found that 1) parity affects both the mean and variance of gestational duration, and 2) clinical history of preterm delivery acts both as pregnancy-specific and shared factor, where the effect of a previous preterm delivery on gestational duration is similar across pregnancies, but the magnitude of effect is pregnancy-specific. Family history of preterm delivery, especially on the mother’s side, reduces the mean of gestational duration and is a shared factor from the second pregnancy. For the first pregnancy a maternal sister delivering preterm is a pregnancy specific factor in the first pregnancy. Our study provides further insights into how parity, a woman’s clinical history of preterm delivery and family history of preterm delivery are acting, which can further aid in clinical risk profiling of at-risk pregnancies and in the design of future epidemiological and genomic studies of the timing of parturition.

## Data Availability

Due to sensitive information, access to raw data requires an application to the National Board of Health and Welfare and Statistics Sweden, conditioned on an approved ethical application (from Swedish Ethical Review Authority gave ethical approval).

## Acknowledgments

Thanks to the National Board of Health and Welfare and Statistics Sweden for providing the data. We would also like to thank our funding support: Sahlgrenska Academy at the University of Gothenburg, The Swedish Research Council, and Swedish government grants to researchers in the public health sector.

## Statements & Declarations

### Author Contributions

Conceptualization: Karin Ytterberg, Pol Sole-Navais, Christopher Flatley, Julius Juodakis; Methodology: Karin Ytterberg, Pol Sole-Navais, Julius Juodakis, Christopher Flatley, Staffan Nilsson; Formal analysis and investigation: Karin Ytterberg; Writing - original draft preparation: Karin Ytterberg; Writing - review and editing: Karin Ytterberg, Pol Sole-Navais, Julius Juodakis, Christopher Flatley, Staffan Nilsson, Bo Jacobsson. The manuscript was approved by all named authors; Funding acquisition: Bo Jacobsson; Supervision: Bo Jacobsson, Staffan Nilsson, Pol Sole-Navais.

### Ethics declarations Competing interests

The authors have no competing interests to disclose.

### Ethical approval

The work was approved by the Swedish Ethical Review Authority (Dnr. 2022-05062-02). The National Board of Health and Welfare permitted the use of the data from the Swedish Medical Birth Register and Statistics Sweden approved the use of the Multigenerational Register and Education Register. All individual-level data are anonymous.

### Informed consent

Informed consent is not required due to the nature of register data.

