## Supplementary Material for "Exploring the impact of parity and its interaction with history of preterm delivery on gestational duration"

### 1. Supplementary Methods

##### Supplementary Methods 1: Generalised Additive Models for Location Scale and Shape (GAMLSS)

The data cleaning for outliers based on deviating child weight and gestational duration correlation was performed with a Generalised Additive Models for Location Scale and Shape (GAMLSS) to fit the non-linear relationship between birth weight and gestational age. A generalised additive model utilizing a Box-Cox-t distribution and cubic spline (cs) smoothing term enabled the calculation of percentile curves and Z-scores derived from these. Any values above 4.5 or below -4.5 standard deviations were considered outliers and were excluded.

##### Supplementary Methods 2: Simulation group effect bias

Two groups of mothers were simulated (n = 20 000 in each group) to investigate the hypothesis of a group effect (that the total number of children a mother will have is associated with gestational duration, possibly due to maternal socio-economic factors). Each mother was assigned a fixed parity effect for parities zero to three (between –0.5 and 0.5 days), a random effect, and a group effect. The random effect was generated randomly from a normal distribution with $\mu = 0$ and $\sigma^2 = 1$. The two groups of mothers were compared given three different scenarios:

1. Every mother in the two groups had an equal number of live births (in total 4) with the group effect set to –1 day.
2. The groups had a different number of children (2 vs 4) but no group effect.
3. The mothers in the different groups had different total numbers of births (2 vs 4) and a group effect set to –1 day (in the 4-child group).

All scenarios was analysed with linear and linear mixed models, with parity as a four-level covariate (reference group parity 0).

#### 2. Supplementary Figures

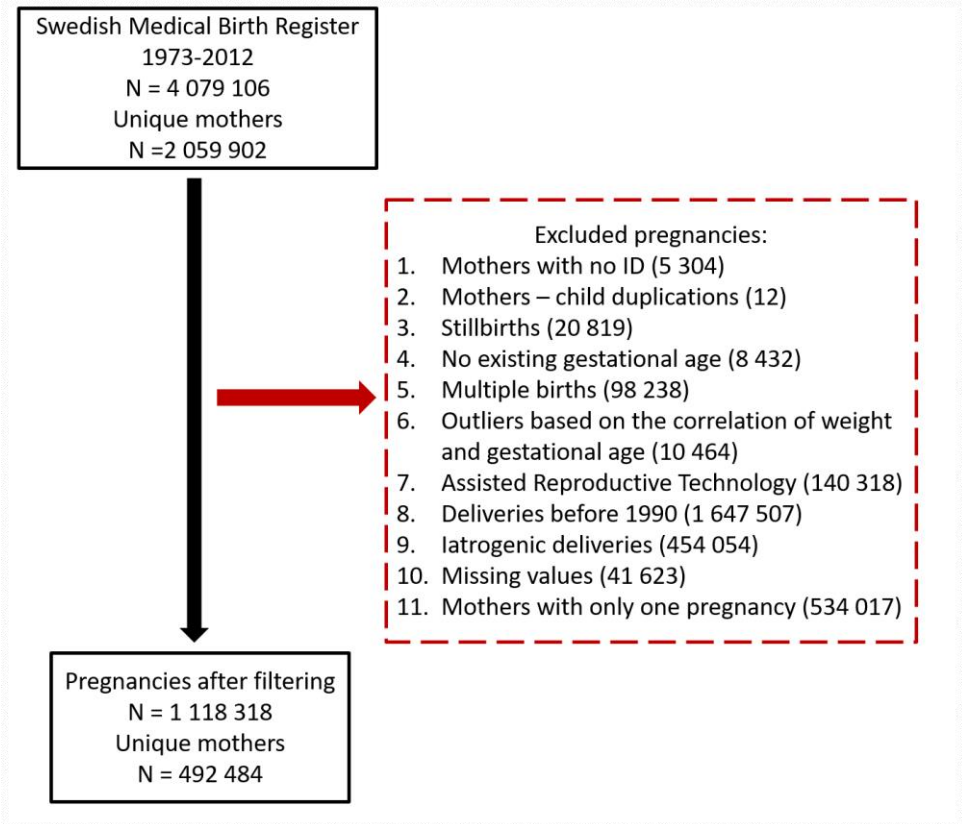

**Supplementary Fig. 1** Flow chart of the stepwise data cleaning process of the Swedish Medical Birth Register. The pregnancies that were excluded during each step of the data cleaning are indicated within parentheses.

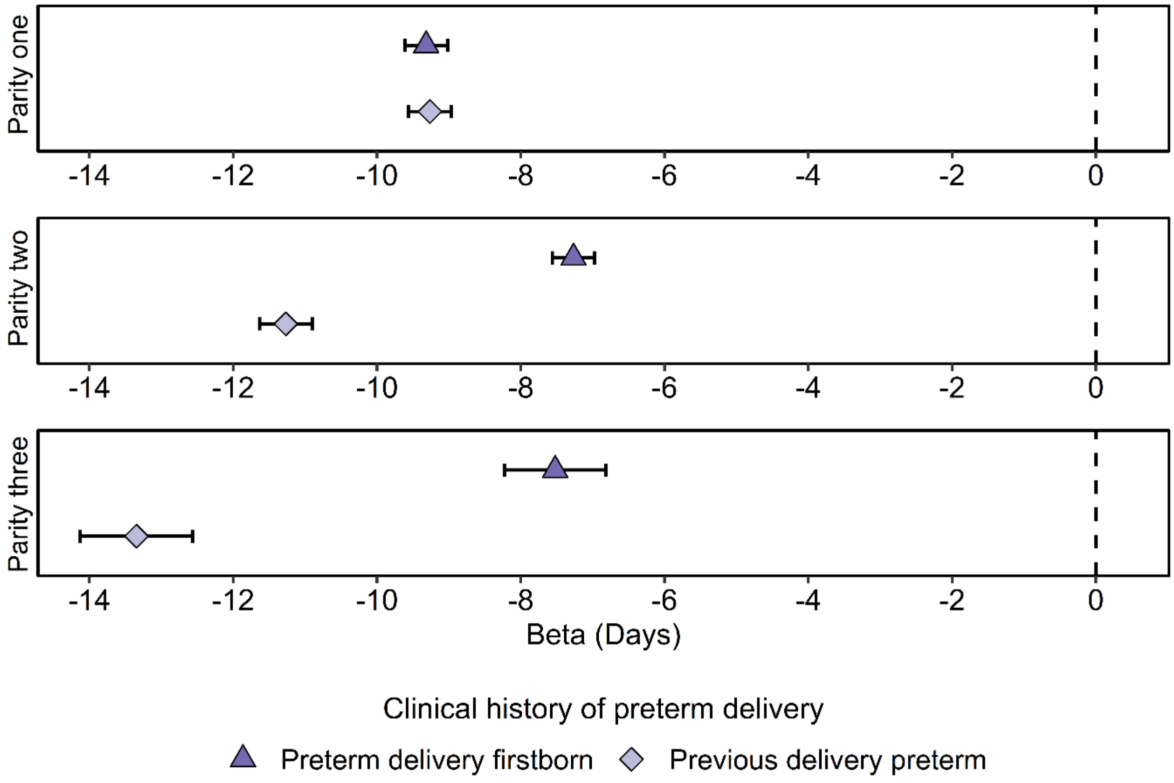

**Supplementary Fig. 2** The modifying effect of a woman's clinical history of spontaneous preterm delivery on gestational duration by parity. The triangle represents the effect on gestational duration in days if a firstborn was spontaneously delivered preterm, while the diamond represents the effect on gestational duration in day if the previous pregnancy was a spontaneous preterm delivery. For the second pregnancy, the two history of preterm delivery measurements have an identical definition. Linear models ran separately for each parity with Set 2 and the specific clinical history of preterm delivery variable. The sample size for each model was: Preterm delivery firstborn: parity one 93 611; parity two 93 071; parity three 17 875, and Previous delivery preterm: parity one 91 720; parity two 100 808; parity three 23 721.

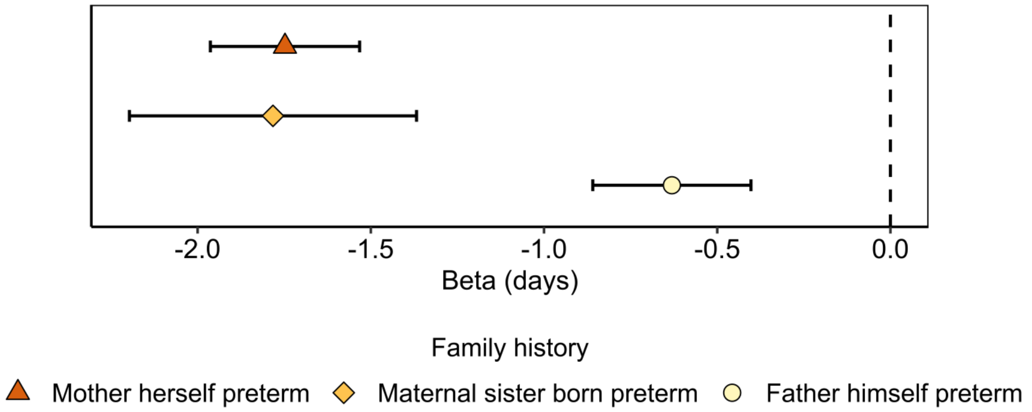

**Supplementary Fig. 3** Effect on gestational duration by family history of preterm delivery. The linear mixed models were run separately for each event of family history; mother herself born preterm (triangle), maternal sister born preterm (diamond), and father born preterm (circle). The sample size for the different models: Mother herself born preterm: 377 101 pregnancies and 168 115 unique mothers, Maternal sibling born preterm: 103 966 pregnancies and 45 735 unique mothers, and Father himself born preterm: 273 912 pregnancies and 130 502 unique mothers. The models were adjusted for Set 2 and the specific family history variable.

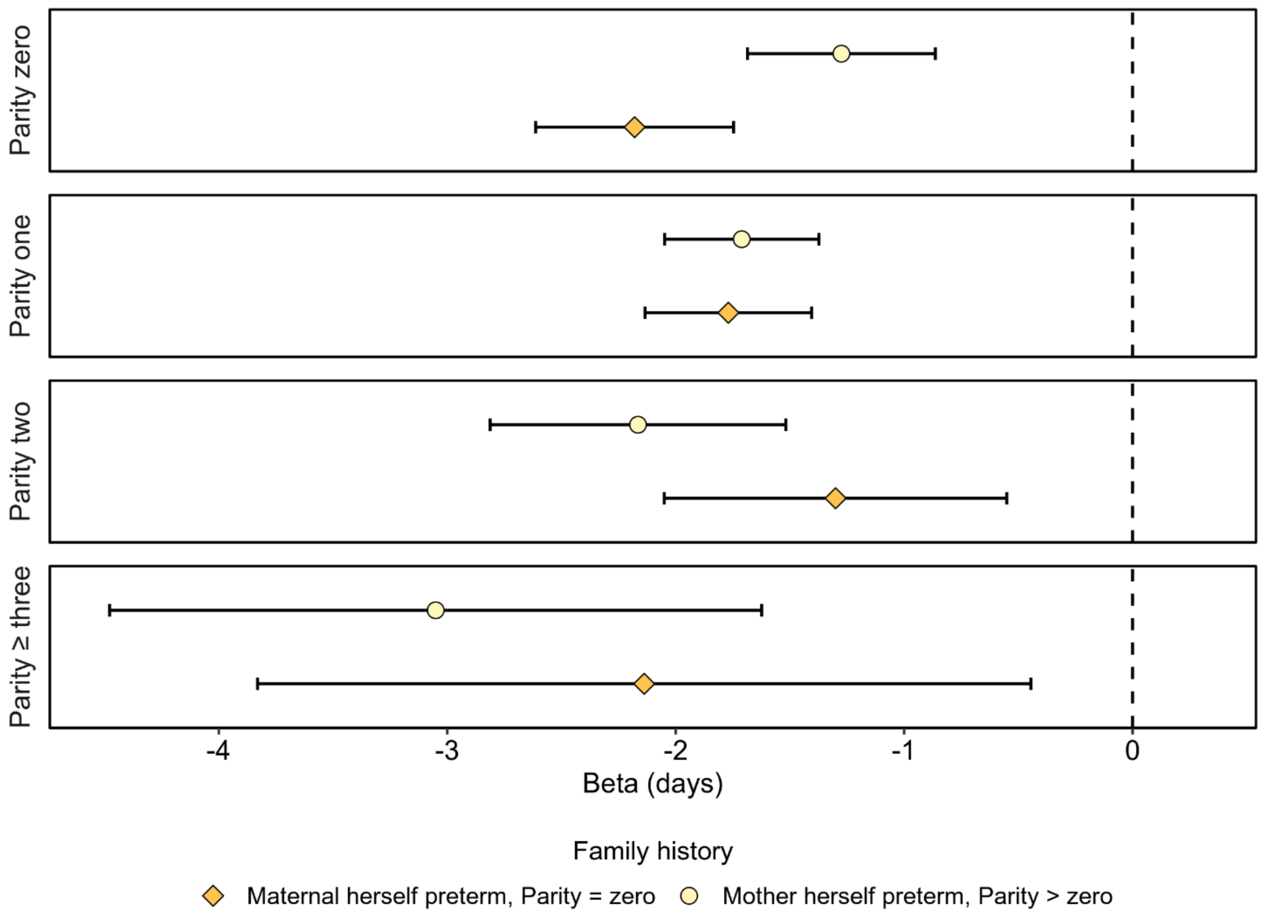

**Supplementary Fig. 4** The effect on gestational duration by the three-way interaction mother herself born preterm, parity and parity of the mother. The figure illustrated the effect on gestational duration in days of a mother being born preterm as a firstborn (diamond) or in any higher (circle) parity. The analysis was performed with linear regressions with Set 2 and the family history variable mother herself born preterm and was stratified by parity and maternal parity. The sample size for each model: Mother herself born preterm, Parity = zero: Parity zero: 67 500; Parity one 68 641; Parity two; 17 804; Parity ≥ 3 3 969, and Mother herself born preterm, Parity ≥ zero: Parity zero 93 460; Parity one 95 167; Parity two 24 927; Parity ≥ 3 5 629.

**(a)**

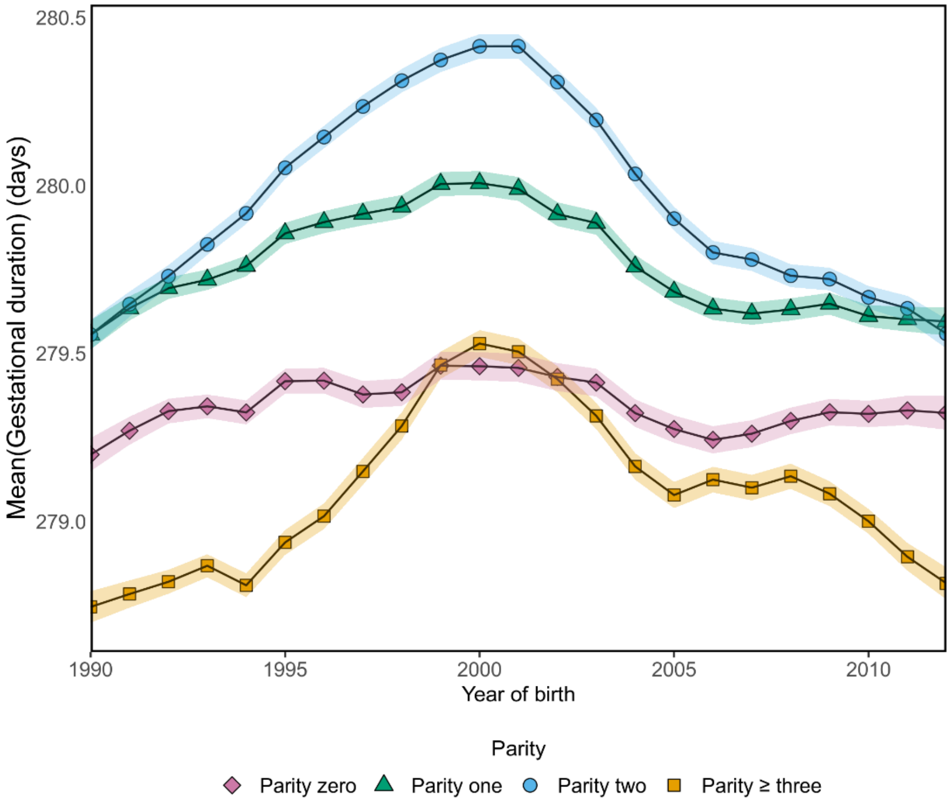

280.5
280.0
279.5
279.0
Mean(Gestational duration) (days)
1990
1995
2000
2005
2010
Year of birth
Parity
Parity zero
Parity one
Parity two
Parity ≥ three

**(b)**

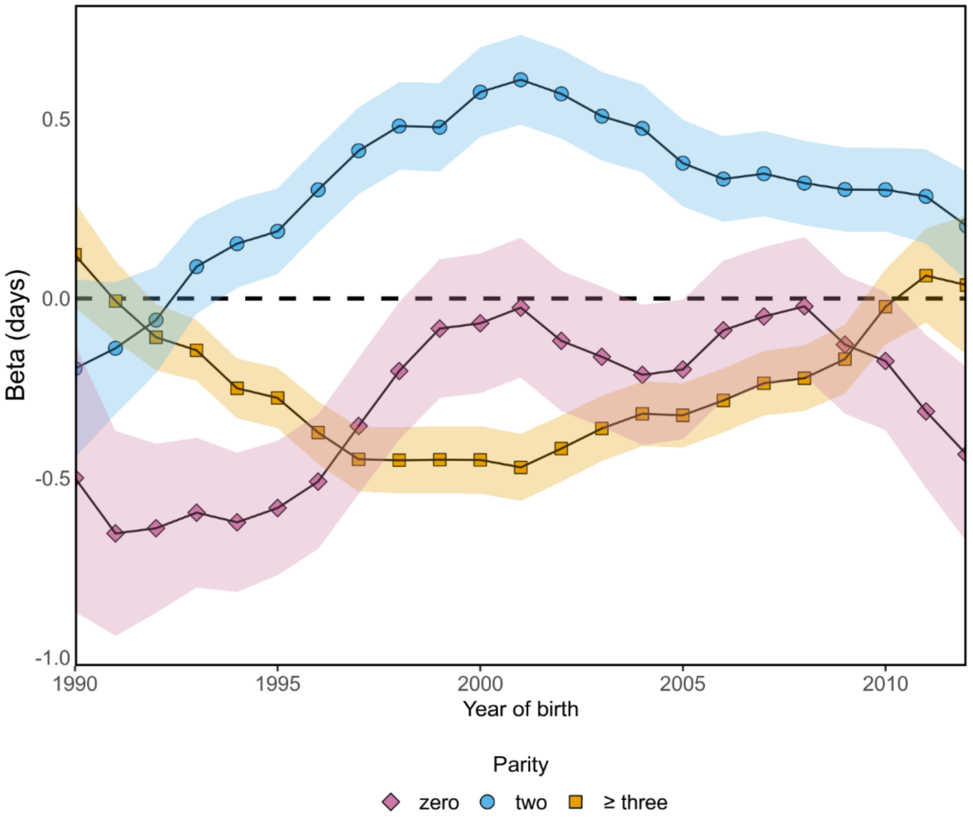

**Supplementary Fig. 5** Interaction of parity and year of birth on gestational duration (1990-2012). (a) The mean gestational duration each year of the cohort by parity (first pregnancy = diamond, second pregnancy = triangle, third pregnancy = circle, and pregnancy four and later = square) (b) The multivariate analyses of gestational duration by parity for each year of birth. The reference group was the second pregnancy (dashed line) and compared to the first (diamond), third pregnancy (circle) and pregnancy four and later (square) in linear mixed models. Each estimate in (a) and (b) were estimated with an interval of +/- 3 years and were adjusted for Set 2 (excluding year of delivery) in linear mixed models. Sample size was varying between 110 289 to 277 606 in the models.

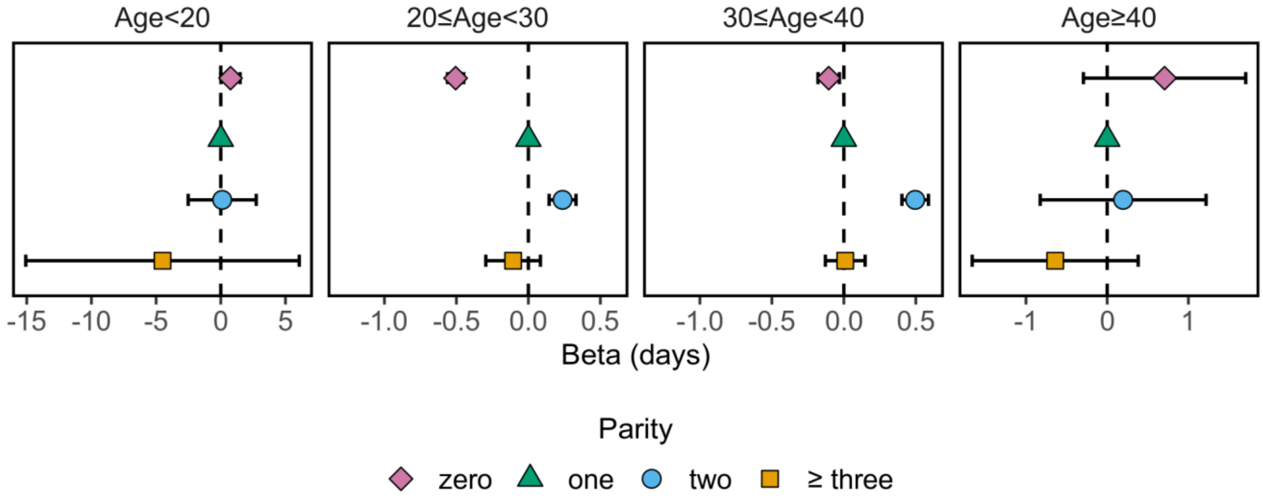

**Supplementary Fig. 6** The modifying effect of maternal age on gestational duration by parity in linear mixed models. The reference group was the second pregnancy (triangle) and compared to the first (diamond), third pregnancy (circle) and pregnancy four and later (square). In each model at least two pregnancies in each mother were required. The models were stratified based on maternal age and adjusted for Set 2 (excluding maternal age). The sample size for each model: Maternal age < 20: 3 538 with 1 743 unique mothers; 20 ≤ Maternal age < 30: 451 653 with 212 151 unique mothers; 30 ≤ Maternal age < 40 320 412 with 151 295 unique mothers; Maternal age ≥ 40: 3 853 with 1 879 unique mothers.

#### 3. Supplementary Tables

**Supplementary Table 1** Additional maternal health characteristics of 801 843 singleton spontaneous live births by parity in Swedish population [a]

| Variable | Parity | | | | |
|---|---|---|---|---|---|
| | 0 | 1 | 2 | ≥ 3 | All |
| **N** (%) | 298 078 (37) | 331 157 (41) | 123 294 (15) | 49 314 (6.2) | 801 843 (100) |
| **Unwilling subfertility** (%) | 7 593 (2.5) | 3 132 (0.95) | 760 (0.62) | 211 (0.43) | 11 696 (1.5) |
| **BMI**, median (interquartile range) | 22.7 (21.2 - 25.9) | 23.2 (20.9 - 25.0) | 23.7 (21.7 - 26.6) | 25.1 (22.5 - 28.6) | 23.1 (21.2 - 25.8) |
| **Smoking**, median (interquartile range) | Non-smoking (Non-smoking – Non-smoking) | Non-smoking (Non-smoking – Non-smoking) | Non-smoking (Non-smoking – Non-smoking) | Non-smoking (Non-smoking – Non-smoking) | Non-smoking (Non-smoking – Non-smoking) |
| **Diabetes** (%) | 1 649 (0.55) | 2 498 (0.75) | 1 263 (1.0) | 836 (1.7) | 6 246 (0.78) |
| **Preeclampsia** (%) | 3 771 (1.3) | 1 297 (0.39) | 470 (0.38) | 239 (0.48) | 5 777 (0.72) |

[a] Description of the additional maternal covariates in Set 3

**Supplementary Table 2** Simulation of group effect bias

| Coefficients from: | Intercept | Parity one | Parity two | Parity three |
|---|---|---|---|---|
| Linear model 1 | -1.02 | 0.498 | 0.839 | 1.03 |
| Linear mixed model 1 | -1.01 | 0.505 | 0.810 | 1.01 |
| Linear model 2 | -0.513 | 0.508 | 0.813 | 1.03 |
| Linear mixed model 2 | 0.505 | 0.505 | 0.806 | 1.01 |
| Linear model 3 | -0.828 | 0.491 | 0.150 | 0.326 |
| Linear mixed model 3 | -1.01 | 0.505 | 0.660 | 0.864 |

Model 1: Every mother in the two groups had an equal number of live births (in total 4) with the group effect.
Model 2: The groups had a different number of children (2 vs 4) but no group effect.
Model 3: The mothers in the different groups had different total numbers of pregnancies (2 vs 4) and a group effect.

**Supplementary Table 3** Quantiles of gestational duration by parity

| Parity | Quantile (days) | | | | |
|---|---|---|---|---|---|
| | 0.10 | 0.25 | 0.50 | 0.75 | 0.90 |
| **0** | 266 | 274 | 281 | 287 | 291 |
| **1** | 269 | 275 | 281 | 286 | 290 |
| **2** | 269 | 275 | 281 | 286 | 291 |
| **≥3** | 267 | 274 | 281 | 286 | 291 |

The sample size: parity zero 462 021; parity one 418 147; parity two 169 453; parity three and higher 68 697

**Supplementary Table 4** Including covariates regarding clinical history of preterm delivery for all previous pregnancies in linear models in a mothers third pregnancy

| Exposures | Estimate (days) | 95% Confidence interval | P-value |
|---|---|---|---|
| Firstborn preterm | -5.57 | -5.79, -5.34 | $< 1 \times 10^{-15}$ |
| Previous delivery preterm | -9.94 | -10.2, -9.63 | $< 1 \times 10^{-15}$ |

**Supplementary Table 5** Including covariates regarding clinical history preterm delivery for all previous pregnancies in linear models in a mothers fourth pregnancy

| Exposures | Estimate (days) | 95% Confidence interval | P-value |
|---|---|---|---|
| Firstborn preterm | -4.57 | -5.06, -4.09 | $< 1 \times 10^{-15}$ |
| Preterm delivery in the second pregnancy | -6.77 | -7.34, -6.19 | $< 1 \times 10^{-15}$ |
| Previous delivery preterm | -11.2 | -11.89, -10.6 | $< 1 \times 10^{-15}$ |
